# Clinical features and progression of acute respiratory distress syndrome in coronavirus disease 2019

**DOI:** 10.1101/2020.02.17.20024166

**Authors:** Yanli Liu, Wenwu Sun, Jia Li, Liangkai Chen, Yujun Wang, Lijuan Zhang, Li Yu

## Abstract

**Background:** The outbreak of severe acute respiratory syndrome coronavirus 2 (SARS-CoV-2) results in a cluster of coronavirus disease 2019 (COVID-19). We reported the clinical characteristics of COVID-19 patients with acute respiratory distress syndrome (ARDS), and further investigated the treatment and progression of ARDS in COVID-19.

**Methods:** This study enrolled 109 patients with COVID-19 admitted to the Central Hospital of Wuhan, a designated hospital in Wuhan, China, from January 2 to February 1, 2020. Patients were followed up to February 12, 2020. The clinical data were collected from the electronic medical records. The differences in the treatment and progression with the time and the severity of ARDS were determined.

**Findings:** Among 109 patients, mean age was 55 years, and 59 patients were male. With a median 15 days (range, 4 to 30 days) follow-up period, 31 patients (28.4%) died, while 78 (71.6%) survived and discharged. Of all patients, 53 (48.6%) developed ARDS. Compared to non-ARDS patients, ARDS patients were elder (mean age, 61 years *vs*. 49 years), and more likely to have the coexistent conditions, including diabetes (20.8% *vs*. 1.8%), cerebrovascular disease (11.3% *vs*. 0%), and chronic kidney disease (15.1% *vs*. 3.6%). Compared to mild ARDS patients, those with moderate and severe ARDS had higher mortality rates. No significant effect of antivirus, glucocorticoid, or immunoglobulin treatment on survival was observed in patients with ARDS.

**Interpretation:** The mortality rate increased with the severity of ARDS in COVID-19, and the effects of current therapies on the survival for these patients were not satisfactory, which needs more attention from clinicians.

**Funding:** Health and Family Planning Commission of Wuhan Municipality.

## Research in context

### Evidence before this study

We searched PubMed and the China National Knowledge Infrastructure database for articles published up to Feb 24, 2020, using the keywords “novel coronavirus”, “2019 novel coronavirus”, “2019-nCoV”, “SARS-CoV-2”, “COVID-19”, “pneumonia”, “coronavirus”, AND “clinical feature”, “mortality”, AND “acute respiratory distress syndrome”, “ARDS”, for articles published in both Chinese and English. We found several recent articles describing the clinical characteristics of COVID-19 patients. One autopsy report described pathological findings of a 50-year-old COVID-19 patient with ARDS. A recent research with 52 critically ill patients published in *The Lancet Respiratory Medicine* indicated that older patients concurrent ARDS are at increased risk of death. No published work about the comprehensive description of clinical features, treatment, and mortality according to the severity of ARDS in COVID-19 patients.

### Added value of this study

We report the differences in the clinical manifestations between COVID-19 patients with and without ARDS. Among 109 patients, 53 (48.6%) of them developed ARDS. Compared with non-ARDS patients, patients with ARDS were elder and more likely to have coexistent diseases. The mortality rate in ARDS patients (49.1%) was significantly higher than that in non-ARDS patients (8.9%). The clinical characteristics of COVID-19 patients varied with the severity of ARDS. High mortality rates were found in patients with moderate and severe ARDS. The survival of COVID-19 patients with ARDS was not significantly improved by the antivirus, glucocorticoid, or immunoglobulin treatment.

### Implications of all the available evidence

In the front-line epidemic area, COVID-19 patients with moderate-to-severe ARDS had high mortality rates. The current medical treatments might not have a satisfactory effect on the in-hospital survival of COVID-19 patients with ARDS. For clinicians, it is necessary to pay close attention to COVID-19 patients who are at high risk for ARDS. The risk stratification and therapeutic strategy for COVID-19 patients should be tailored according the variations in ARDS severity. It is essential for intensive care physicians to participate in treatment decision-making and management in the early stages of the COVID-19 outbreak.

## Introduction

The emergent outbreaks of coronavirus disease 2019 (COVID-19) caused by the novel severe acute respiratory syndrome coronavirus 2 (SARS-CoV-2) remain a threat to the public health worldwide ^1,2^. As of February 24, 2020, 77,262 cases were confirmed, and 2,595 death cases were recorded in China. The number of confirmed cases in other countries are ascending as well ^3-5^. COVID-19 may impose a great socioeconomic, public health, and clinical burden, especially in the low-income and middle-income countries.

Growing studies identified the clinical features of COVID-19 ^6-8^. Similar to SARS in 2003, this infectious disease results in the high possibility of the admission to the intensive care unit (ICU) and the mortality ^6^. Notably, due to the cytokine cascade within a short period, the critically ill patients with COVID-19 were more likely to develop acute respiratory distress syndrome (ARDS) and receive oxygen treatment ^6^. ARDS was the most common complication in COVID-19 patients, with a high mortality rate ^6,7^. The latest publication in *The Lancet Respiratory Medicine*, Yang *et al*. ^9^ reported that 67% of critically ill patients develop ARDS. However, little is known regarding the clinical characteristics, treatment, and progression of COVID-19 patients according to the severity of ARDS.

In this study, we retrospectively reviewed the clinical data of patients with COVID-19 who were admitted to the Central Hospital of Wuhan, and determined the differences in the clinical characteristics between COVID-19 patients with and without ARDS. Additionally, to reveal the evolvement of ARDS in COVID-19, we also investigated the clinical features and therapies of COVID-19 patients at each severity level of ARDS. Our study might provide new insight into the risk stratification and therapeutic strategy for COVID-19 patients.

## Methods

### Study Population

We retrospectively analyzed the data of confirmed COVID-19 patients admitted to the Central Hospital of Wuhan between January 2 and February 1, 2020. The Central Hospital of Wuhan is one of the first designated hospitals to receive patients with COVID-19. According to the World Health Organization interim guidance ^10^, the diagnostic criteria of COVID-19 was based on the virus RNA detection, the clinical characteristics, the chest imaging, and the ruling out common pathogen. Patients with malignant tumors, previous craniocerebral operation, or died on admission were excluded. In this study, we also excluded patients who had been transferred to other hospitals for advanced life support and patients with mild symptoms who had been transferred to mobile cabin hospitals. The clinical outcomes of patients were followed up to February 12, 2020. Patients who were still in hospital before February 12, 2020 were excluded. Finally, 109 confirmed COVID-19 patients were included in the analyses. All data were anonymous, and the requirement for informed consent was waived. The Ethics Committees of the Central Hospital of Wuhan approved this study.

### Data Collection

Case report forms, nursing records, laboratory findings, and radiological features were reviewed. All data were collected onto the standardized forms from the electronic medical records in the hospital. Two senior physicians independently reviewed the data. Information on demographic data, symptoms, underlying comorbidities, chest computed tomographic images, and laboratory results were included. All treatment measures were collected during the hospitalization, such as antiviral therapy, antibacterial therapy, corticosteroid therapy, immune support therapy, and respiratory support. The time of disease onset was defined as the day of symptom onset. The time from the first symptom to the fever clinics, hospital admission, and clinical outcomes were recorded. A confirmed respiratory tract specimen was defined as positive for SARS-CoV-2. Repeated tests for SARS-CoV-2 were done in the confirmed patients to show viral clearance before the hospital discharge. The Berlin definition was applied to determine the presence and severity of ARDS ^11^. Acute Physiology and Chronic Health Evaluation II (APACHE II), Sequential Organ Failure Assessment (SOFA) scores and CURB-65 criteria ^12^ were determined within 24 hours after admission.

### Detection of Coronavirus by Real-Time Reverse Transcription Polymerase Chain Reaction

Throat swab samples were collected from the suspected patients, and the method of virus RNA detection was reported ^7^. Briefly, the presence of SARS-CoV-2 in the respiratory specimens was detected by real-time reverse transcription polymerase chain reaction (RT-PCR). Two target genes were used, including open reading frame lab (ORF1ab) and nucleocapsid protein (N), and the sequences were as follows: ORF1ab: forward primer CCCTGTGGGTTTTACACTTAA; reverse primer ACGATTGTGCATCAGCTGA. N: forward primer GGGGAACTTCTCCTGCTAGAAT; reverse primer CAGACATTTTGCTCTCAAGCTG. The real-time RT-PCR assay was conducted in line with the manufacturer’s protocol (Beijing Genomics Institution and Geneodx biotechnology Co. Ltd). Positive test results for two target genes were considered as laboratory-confirmed infection.

### Statistical Analysis

Normally distributed continuous variables were presented as mean ± SD, compared by Student’s t-test. Skew distributed continuous variables were shown as median (interquartile range [IQR]), analyzed by Mann–Whitney test or Kruskal–Wallis test. Categorical variables were compared by Chi-square test or the Fisher’s exact test. Kaplan–Meier methods were used for survival plotting, and log-rank test for comparison of survival curves. The dynamic trajectory in laboratory parameters was plotted using GraphPad Prism 8 (GraphPad Software, Inc). Generalized linear mixed models examined the differences in the laboratory data between ARDS and non-ARDS groups over time. All statistical analysis were performed by the statistical software packages R (http://www.R-project.org, The R Foundation) and the EmpowerStats (http://www.empowerstats.com, X&Y Solutions, Inc., Boston, MA) with a two-sided significance threshold of *P* <0.05.

## Results

### Clinical Characteristics

A total of 109 patients with COVID-19 were included in the current study. The baseline clinical parameters of overall COVID-19 patients and COVID-19 patients with and without ARDS were summarized in **Table 1**. Mean age of all subjects was 55 years (IQR, 43-66 years; range, 22-94 years), and 54.1% of all patients were male. The most common symptom at the illness onset was fever (82.6%), followed by dry cough (61.5%) and fatigue (56.9%). Seventy-six patients (69.7%) had underlying comorbidities, including hypertension (33.9%), diabetes (11.0%), and chronic kidney disease (9.2%). The first virus RNA detection rate was 24.8% in the fever clinics, and all patients were reconfirmed by repeated virus RNA testing after admission. Among 109 patients, 53 (48.6%) of them developed ARDS, 100 (91.7%) of them had bilateral involvement of the chest radiographs, and 28 (25.7%) patients received high-flow nasal oxygen ventilation, while 31 (28.4%) died.

**Table 1.** Baseline clinical features of subjects with COVID-19.

|  | No. (%) |  |  | *P*-value<sup>a</sup> |
| --- | --- | --- | --- | --- |
|  | Total (*n* = 109) | Non-ARDS (*n* = 56) | ARDS (*n* = 53) |  |
| **Age, median (IQR), y** | 55 (43-66) | 49 (37-59) | 61 (52-70) | **<0.001** |
| **Male** | 59 (54.1) | 31 (55.4) | 28 (52.8) | 0.79 |
| **Entering complaint** |  |  |  |  |
| Fever | 90 (82.6) | 48 (85.7) | 42 (79.2) | 0.37 |
| Dry cough | 67 (61.5) | 36 (64.3) | 31 (58.5) | 0.53 |
| Fatigue | 62 (56.9) | 26 (46.4) | 36 (67.9) | **0.02** |
| Stethalgia | 7 (6.4) | 4 (7.1) | 3 (5.7) | 0.75 |
| Diarrhea | 12 (11.0) | 6 (10.7) | 6 (11.3) | 0.92 |
| **Comorbidities** |  |  |  |  |
| COPD | 4 (3.7) | 2 (3.6) | 2 (3.8) | >0.99 |
| Hypertension | 37 (33.9) | 16 (28.6) | 21 (39.6) | 0.22 |
| Diabetes | 12 (11.0) | 1 (1.8) | 11 (20.8) | **0.002** |
| Cardiovascular disease | 7 (6.4) | 4 (7.1) | 3 (5.7) | >0.99 |
| Cerebrovascular disease | 6 (5.5) | 0 (0.0) | 6 (11.3) | **0.01** |
| Chronic kidney disease | 10 (9.2) | 2 (3.6) | 8 (15.1) | **0.049** |
| **Onset of symptom to fever clinics, median (IQR), d** | 4 (2-6) | 4 (2-7) | 4 (2-6) | 0.88 |
| **Onset of symptom to hospital admission, median (IQR), d** | 7 (5-9) | 7 (5-9) | 7 (5-9) | 0.58 |
| **Hospital stays, median (IQR), d** | 15 (11-24) | 15 (9-27) | 16 (11-23) | 0.41 |
| **Nucleic acid test (+) in fever clinics** | 27 (24.8) | 12 (21.4) | 15 (28.3) | 0.41 |
| **Bilateral involvement of chest radiographs** | 100 (91.7) | 47 (83.9) | 53 (100) | **0.002** |
| **Fever clinics treatment** |  |  |  |  |
| Antibiotics | 105 (96.3) | 55 (98.2) | 50 (94.3) | 0.35 |
| Antivirus | 105 (96.3) | 55 (98.2) | 50 (94.3) | 0.35 |
| Glucocorticoid therapy | 43 (39.4) | 20 (35.7) | 23 (43.4) | 0.41 |
| Intravenous immunoglobulin | 32 (29.4) | 19 (33.9) | 13 (24.5) | 0.28 |
| **In-patient treatment** |  |  |  |  |
| Antibiotics | 106 (97.2) | 53 (94.6) | 53 (100.0) | 0.24 |
| Antivirus | 107 (98.2) | 55 (98.2) | 52 (98.1) | >0.99 |
| Glucocorticoid therapy | 72 (66.1) | 35 (62.5) | 37 (69.8) | 0.42 |
| Intravenous immunoglobulin | 61 (56.0) | 32 (57.1) | 29 (54.7) | 0.80 |
| **High-flow nasal ventilation** | 28 (25.7) | 2 (3.6) | 26 (49.1) | **<0.001** |
| **Death** | 31 (28.4) | 5 (8.9) | 26 (49.1) | **<0.001** |

Abbreviations: ARDS, acute respiratory distress syndrome; COPD, chronic obstructive pulmonary disease; IQR, interquartile range;
<sup>a</sup> *p* values indicate differences between non-ARDS and ARDS patients.

Compared with non-ARDS patients with COVID-19, ARDS patients were elder (mean age, 61 years *vs*. 49 years), and more likely to have coexistent diabetes (20.8% *vs*. 1.8%), cerebrovascular disease (11.3% *vs*. 0%), and chronic kidney disease (15.1% *vs*. 3.6%). The bilateral involvement in the chest radiographs was identified in all ARDS patients, but in 83.9% non-ARDS patients. The likelihood to receive high-flow nasal ventilation for ARDS patients (49.1%) was significantly higher than that for non-ARDS patients (3.6%). The mortality rate in ARDS patients (49.1%) was also significantly higher than that in non-ARDS patients (8.9%).

### Disease Severity Scores and Laboratory Findings in COVID-19 Patients With and Without ARDS

The disease severity evaluation and laboratory examination of non-ARDS and ARDS patients with COVID-19 on admission were shown in **Table 2**. Compared with non-ARDS subjects, ARDS subjects had significantly higher disease severity on the day of hospital admission (all *P* values <0.001 for CURB-65, SOFA, and APACHE II). ARDS patients were more likely accompanied by lymphopenia on admission (*P* value <0.001). Besides, the levels of lactate, neutrophil count, C-reactive protein (CRP), and procalcitonin were significantly higher in ARDS patients than in non-ARDS patients (median lactate level, 1.6 *vs*. 1.1 mmol/L; median neutrophil count level, 4.1 *vs*. 3.2 ×10^9^/L; median CRP level, 4.9 *vs*. 2.0 mg/dL; median procalcitonin level, 0.15 *vs*. 0.06 ng/mL). Significant increased levels of blood urea nitrogen, serum fibrinogen, D-dimer, and lactate dehydrogenase in ARDS patients were observed, compared with those in non-ARDS patients (all *P* values <0.05).

**Table 2.**
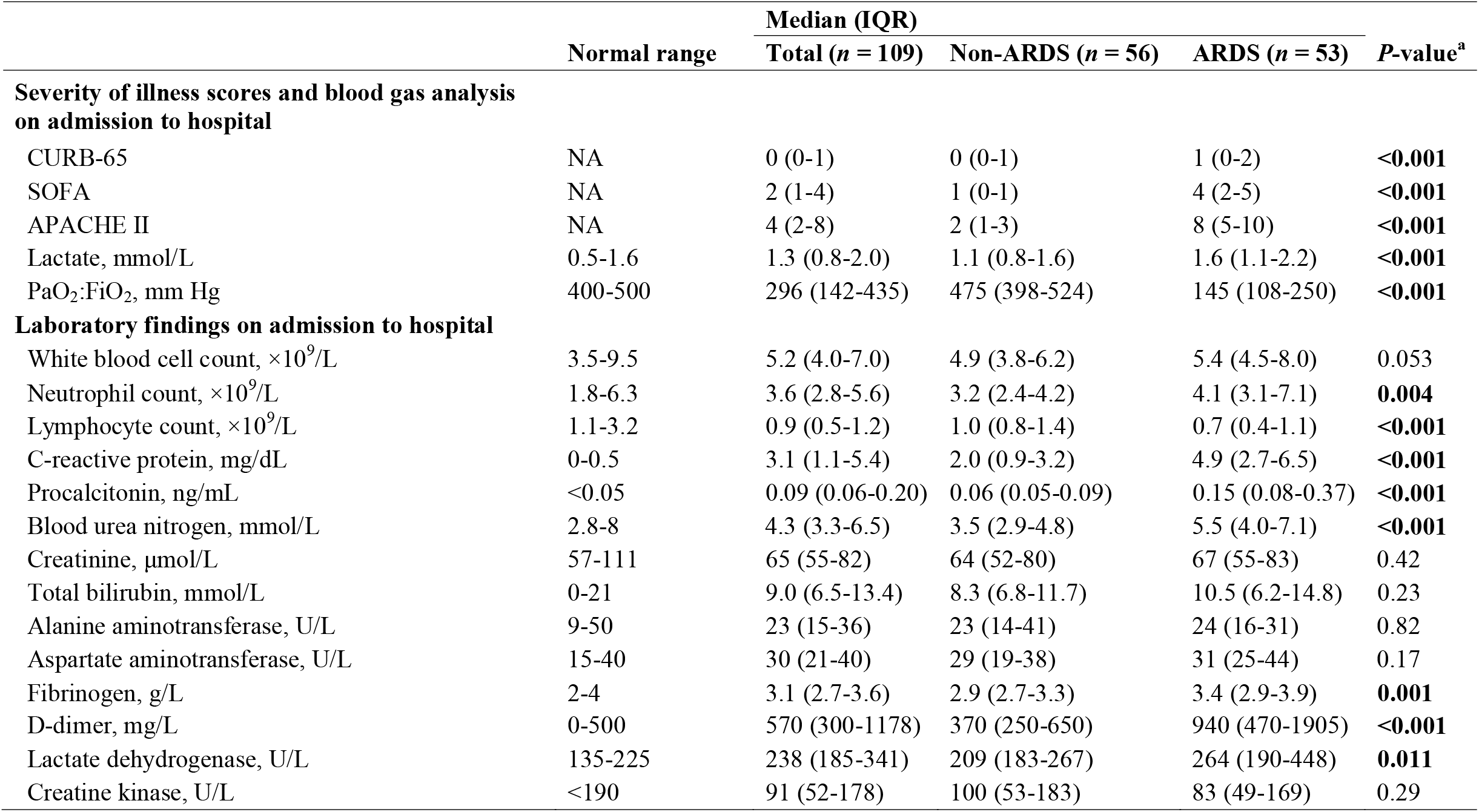
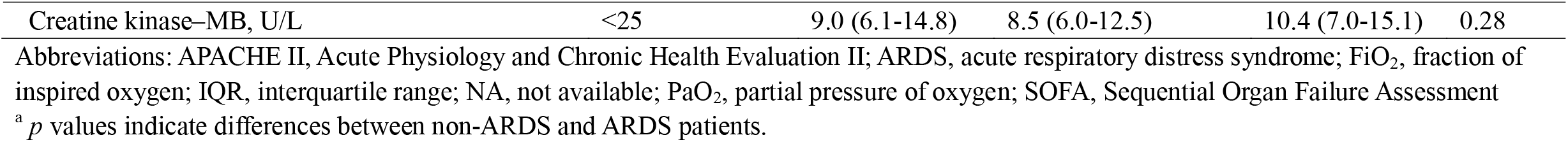
Severity of COVID-19 and laboratory tests in Non-ARDS and ARDS subjects.

As shown in **Figure 1**, the dynamic trajectory in nine laboratory parameters were tracked on Day 1, 3, 7 and 14, respectively. Significant differences in the levels of neutrophil count, lymphocyte count, CRP, procalcitonin, and D-dimer were found between ARDS and non-ARDS patients with COVID-19 (all *P* values <0.05). From Day 1 to 14, ARDS patients had significantly lower lymphocyte count levels, while the levels of other laboratory findings were higher in ARDS patients.

**Figure 1.**
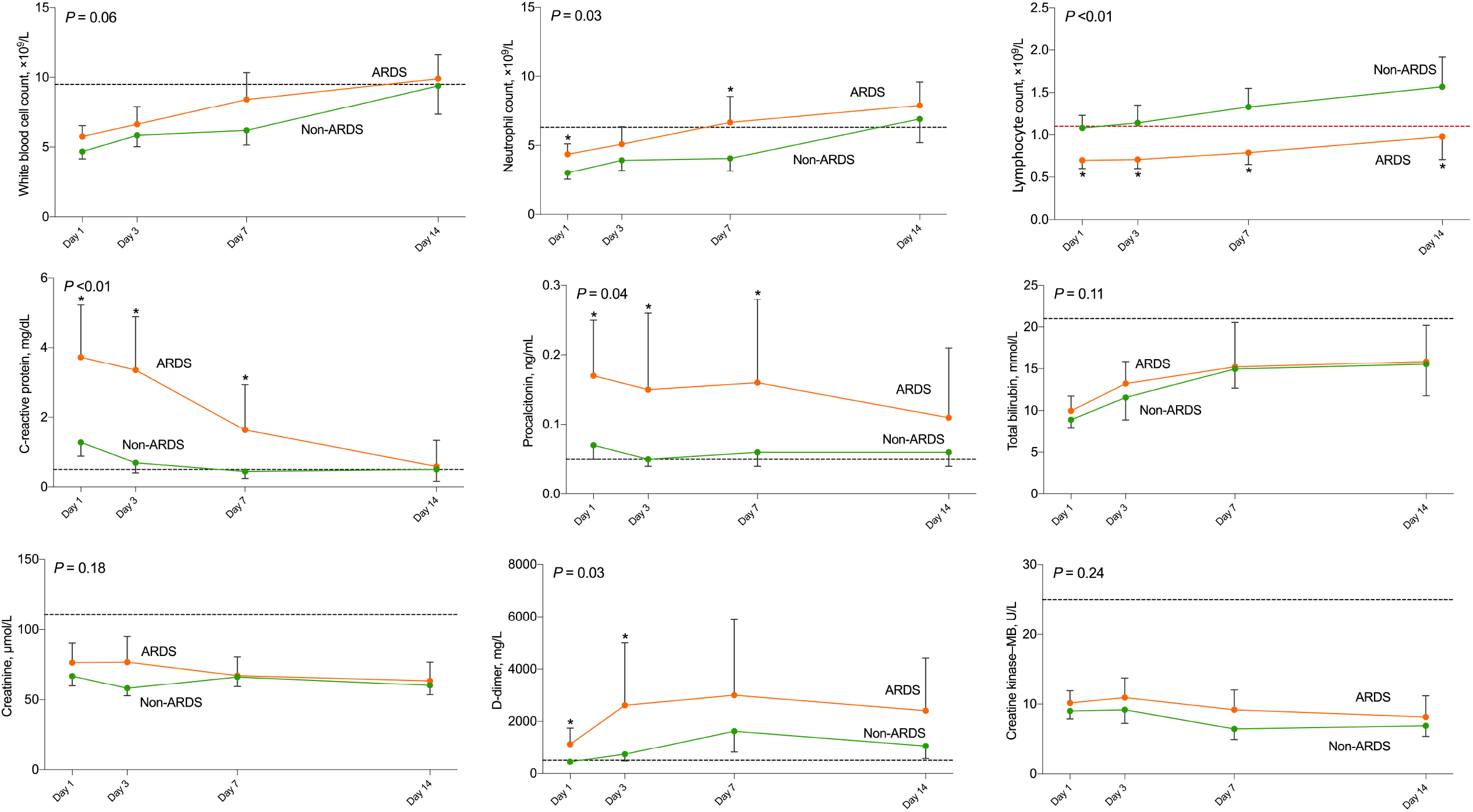
Timeline charts illustrate the laboratory parameters in 109 patients with COVID-19 (56 non-ARDS and 53 ARDS) on day 1, day 3, day 7, and day 14 after admission. Data are represented as geometric mean and 95% confidence interval (one group only shows the upper or lower bar). The dash lines in black show the upper normal limit of each parameter, and the dash line in red shows the lower normal limit of lymphocyte count. * *P* <0.05 for non-ARDS *vs*. ARDS.

### Clinical Profile and Progression of COVID-19 Patients With ARDS

According to the severity of ARDS, we further analyzed the clinical features, treatment, and progression of 53 COVID-19 patients with ARDS. As presented in **Table 3**, the increase in APACHE II, SOFA, and CURB criteria scores occurred concomitantly with the severity of ARDS (all *P* values <0.001). The levels of lactate, blood urea nitrogen, D-dimer, and lactate dehydrogenase ascended with the severity of ARDS (all *P* values <0.05), whereas the lymphocyte count levels decreased (*P* value = 0.045). Patients with moderate-to-severe ARDS were the most likely to receive glucocorticoid therapy (*P* value = 0.02) and high-flow nasal oxygen ventilation (*P* value <0.001). Compared to patients with mild ARDS, those with moderate and severe ARDS had higher mortality rates (*P* value <0.001). Subsequent survival analysis was shown in **Figure 2**. We found no significant effect of antivirus, glucocorticoid, or immunoglobulin treatment on survival in COVID-19 patients with ARDS (all log-rank tests *P* >0.05).

**Table 3.**
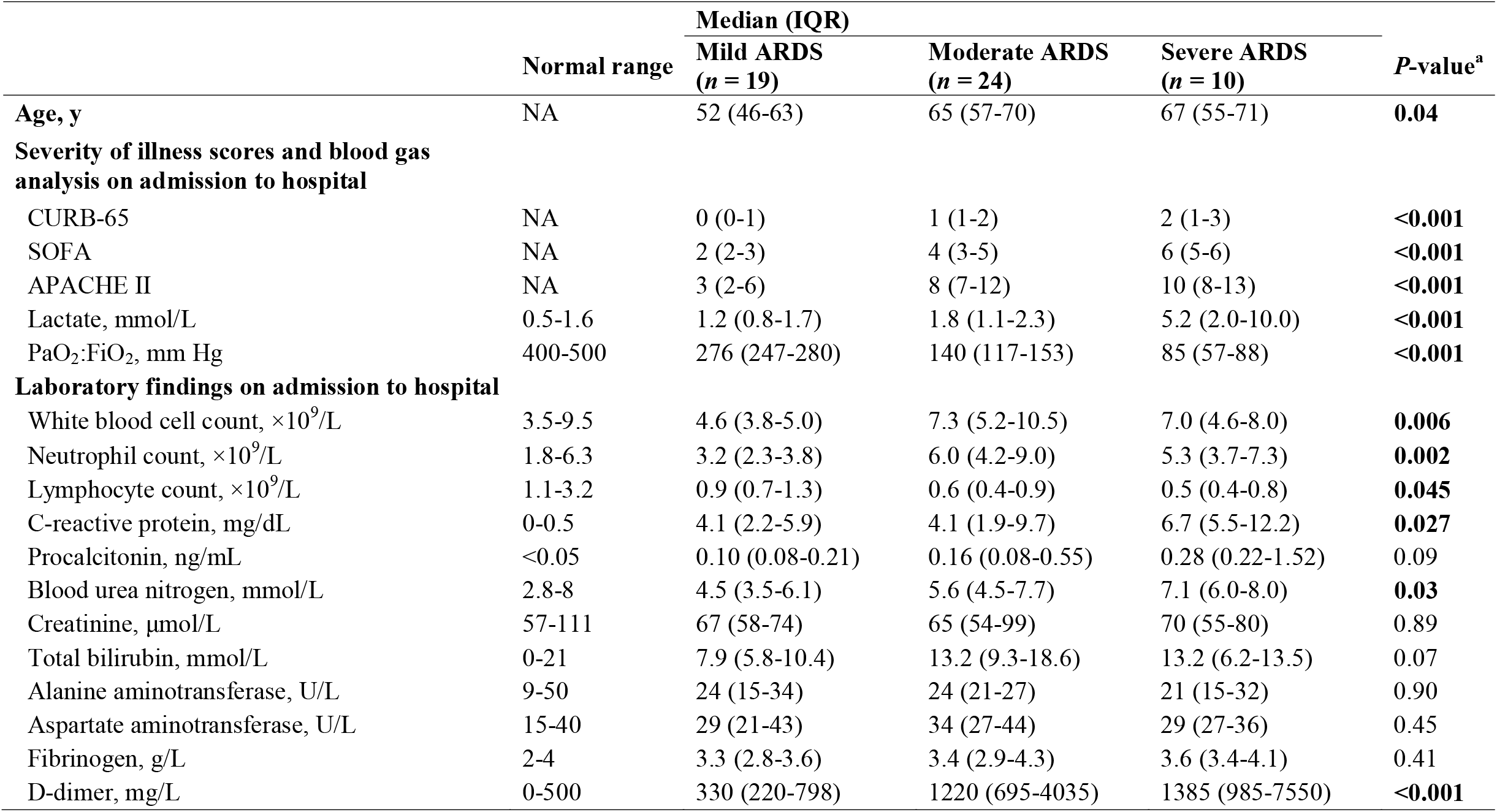
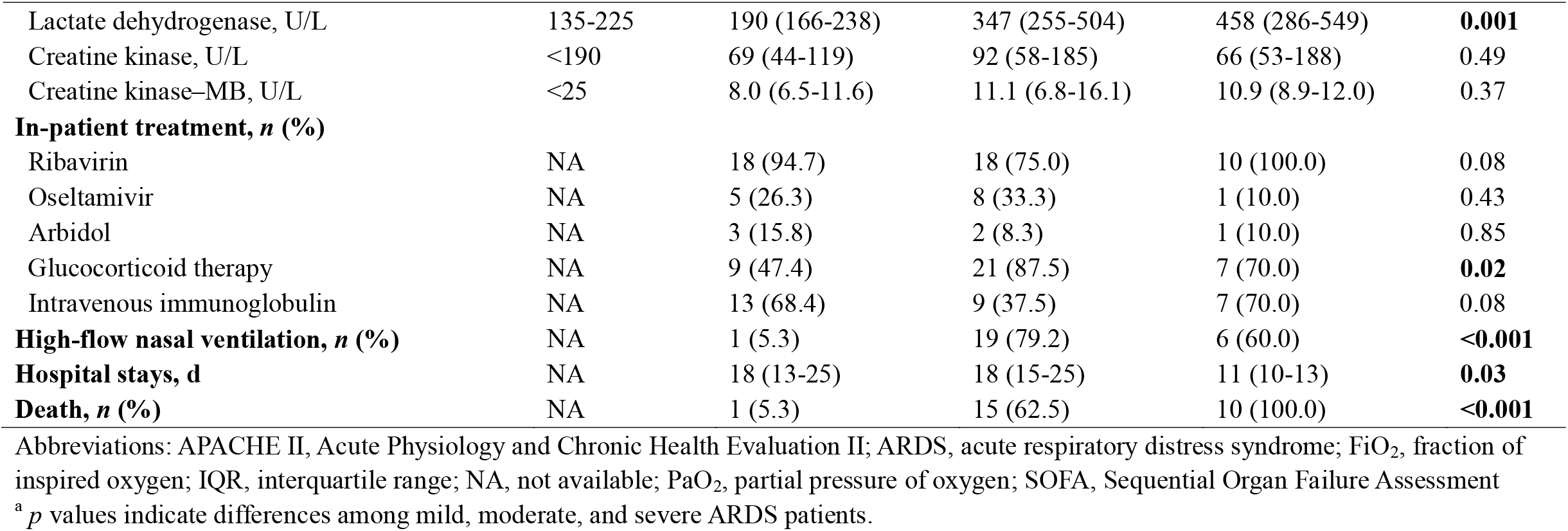
Clinical characteristics, laboratory findings, and treatment according to the severity of ARDS.

**Figure 2.**
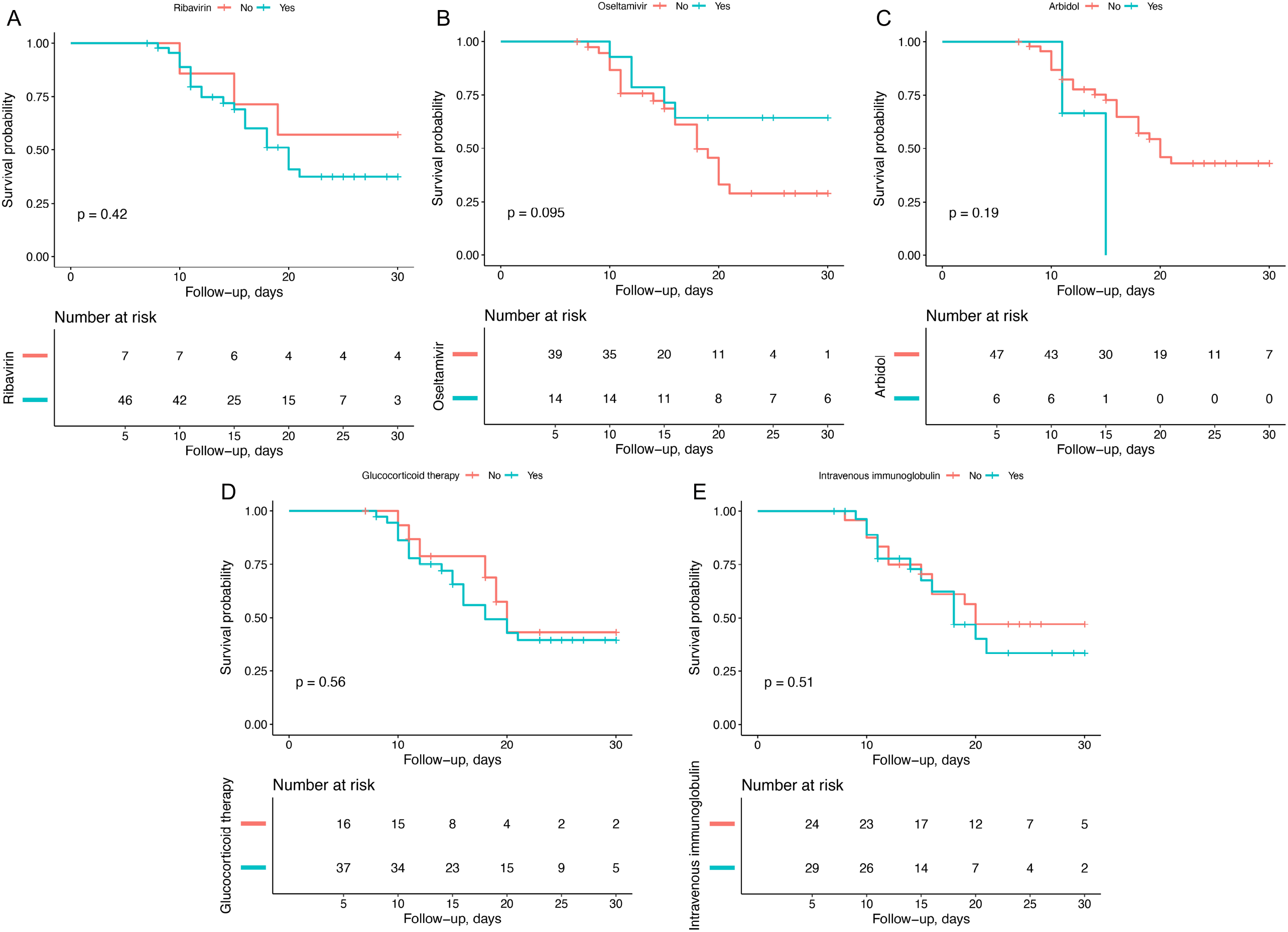
Kaplan-Meier survival curves for 53 COVID-19 patients concurrent ARDS. (A) Ribavirin; (B) Oseltamivir; (C) Arbidol; (D) Glucocorticoid therapy; (E) Intravenous immunoglobulin.

## Discussion

In the present study of ARDS progression in COVID-19, we firstly identified the differences in the clinical manifestations between COVID-19 patients with and without ARDS. Second, the clinical characteristics of COVID-19 patients varied with the severity of ARDS. High mortality rates were observed in patients with moderate-to-severe ARDS. Third, we did not observe any apparent effect of the current therapeutic interventions on the in-hospital survival of COVID-19 patients with ARDS. Our findings might highlight the clinical significance in the enhanced attention towards COVID-19 patients at high risks of ARDS.

To the best of our knowledge, this study is the first to investigate the clinical features and progression of SARS-CoV-2 infected patients according to the severity of ARDS. We revealed that elder patients, or complicated by diabetes, cerebrovascular disease, and chronic kidney disease, were more likely to develop ARDS. This finding might further explain the significant association of age and comorbidity conditions with the poor clinical outcomes in a previous COVID-19 study ^7^. In addition, the elevated levels of neutrophil count, CRP, procalcitonin and D-dimer, but the decreased lymphocyte count levels, were detected in patients with ARDS, rather than those without ARDS. Similarly, the levels of lactate and D-dimer increased with the progression of ARDS due to SARS-CoV-2, while severer lymphopenia occurred over time. Our investigations into these laboratory indicators were comparable with other COVID-19 studies ^6,7,13^, suggesting that cytokine cascade, excessive inflammatory reaction, and coagulation dysfunction might evolve in the course of ARDS resulted from SARS-CoV-2.

Our understanding of this ARDS progression could be furthered by a recent autopsy report on a 50-year-old COVID-19 patient with ARDS ^14^, which was highlighted by the inflammatory infiltration of lymphocytes in both lungs and the immune hyperactivation in the patient. Interestingly, no obvious heart injury occurred in this patient, while the liver injury might be caused by SARS-CoV-2 infection or medication use ^14^. These pathological results were concordant with our clinical observation of a dysregulated inflammatory and immune state in COVID-19 patients with ARDS, which might in turn help to guide our medical therapy.

The medical management of ARDS has advanced remarkably during the past decades^15^. However, the general mortality rates of ARDS caused by all etiologies fluctuated from 11% to 87% ^16^. In our study, most patients with COVID-19 received antibiotics and antivirus treatment, while over half of them were given by glucocorticoid and intravenous immunoglobulin therapies. Moderate-to-severe ARDS patients were more likely to use high-flow nasal oxygen ventilation. Unfortunately, the mortality rate was extremely high in severe ARDS patients with COVID-19, and the survival of COVID-19 patients with ARDS was not improved by the antivirus, glucocorticoid, or immunoglobulin treatment. Some potential reasons might interpret the high mortality we observed: First, due to the SARS-CoV-2 outbreak within a short period, the designated hospitals were overloaded in short term, with inadequate medical facilities and insufficient medical staff. Notably, intensive care resources are in short supply and may fail to meet demand under the increasing number critically ill patients. Second, the Central Hospital of Wuhan is close to Huanan Seafood Market, and the inpatients were the first or the second generation infected patients. In addition, the inadequate preparation and insufficient understanding of the disease might contribute to increased mortality in the front-line epidemic area during the early outbreak period.

The epidemic spread knows no borders. We are deeply concerned about the current outbreak of SARS-CoV-2. As the low-income and middle-income countries are more vulnerable to the lack of medical resources, the mortality rate of infectious diseases may be even higher. Therefore, from a policy perspective, the government should provide capital, technology, and manpower to curb the epidemic. Meanwhile, in the front-line epidemic area, limited medical resources should be allocated reasonably and effectively. Based on our observation, some patients progress quickly to severe pneumonia or ARDS without warning sign. Adequate critical care resources and the involvement of intensive care physicians are essential in the early stages of the SARS-CoV-2 outbreak.

The present study is subjected to several limitations. Due to the retrospective nature of the present study, a systematic selection bias could be introduced. We could not completely address the residual confounding factors as well. Although our laboratory observations showed the significant changes with the time and the severity of ARDS, the clinical predictive value remained to be determined. In addition, our results on the effect of the current treatments should be considered with caution, and high-quality clinical interventional studies are needed. Meanwhile, the benefits of the invasive ventilation therapy on the disease prognosis should be further investigated. In the future, a multi-center and follow-up study with a larger cohort is eagerly warranted.

## Conclusions

In the current study from one of the designated hospitals in Wuhan, China, the significant changes with the time and the severity of ARDS were observed in COVID-19 patients. Moreover, COVID-19 patients with moderate and severe ARDS had high mortality rates. The current medical treatments might not have a significant effect on the in-hospital survival of COVID-19 patients with ARDS. Our observations might help to guide the risk stratification and therapeutic strategy for COVID-19 patients.

## Data Availability

With the permission of the corresponding authors, we can provide participant data without names and identifiers, but not the study protocol, statistical analysis plan, or informed consent form. Data can be provided after the article is published.

## Acknowledgement

We deeply regret and mourn all the lives lost in this disaster of SARS-CoV-2, including our dearest colleague Dr. Wenliang Li. We would like to express our deepest respect to all the people who are currently fighting against the outbreak of COVID-19.

## Authors Contributions

LC and YL had full access to all the data in the study, and took responsibility for the integrity of the data and the accuracy of the data analysis. YL, WS, LC, and LY made substantial contributions to the study concept and design. YL and WS took responsibility for obtaining ethical approval, collecting samples, and confirming data accuracy. LC was in charge of the statistical analysis. YL, JL, and LC were in charge of the manuscript draft. JL, LC, LZ, YW, and LY contributed to critical revision of the report. All authors reviewed and approved the final version.

## Data sharing

The data that support the findings of this study are available from the corresponding authors on reasonable request. We can provide participant data without names and identifiers, but not the study protocol, or statistical analysis plan. After publication of study findings, the data will be available for others to request. Once the data can be made public, the research team will provide an email address for communication. The corresponding authors have the right to decide whether to share the data or not based on the research objectives and plan provided.

